# Identification of common pre-analytical errors during the HIV viral load and EID testing in Cameroon: A call for strengthening the laboratory systems to support follow-up people living with HIV/AIDS

**DOI:** 10.1101/2025.05.05.25327012

**Authors:** Marie Atsama-Amougou, Sabine Ndejo Atsinkou, Yagai Bouba, Hamadou Amadou, Teclaire Elodie Ngo-Malabo, Nadine Nguendjoung Fainguem, Julius Nwobegahay, Comfort Vuchas, Elong Elise, Bertrand Eyoum Bille, David Donchi Nguiala, Emmanuel Biongolo, Boutgam Nadine Lamare, Yakouba Liman, Marcel Tongo, Etienne Mpabuka, Ahidjo Ayouba, Charles Kouanfack, Albert Franck Zeh Meka, Joseph Fokam

## Abstract

**Background:** HIV Viral Load (HIV-VL) and Early Infant Diagnosis (EID) play a pivotal role in the laboratory surveillance, monitoring of HIV/AIDS, and its elimination as a public concern. However, sample rejection resulting from sample non-conformity (SNC) due to inadequate collection, transportation, and management of samples, especially during the pre-analytic phase, negatively affects laboratory performance. This study aimed to characterize errors observed during the pre-analytical phase of HIV-VL and EID testing throughout the different national reference laboratories in Cameroon and factors associated with reasons for rejection.

**Methods:** A descriptive and quantitative study of the non-conformities (NC) identified were collected from January to December 2024 in seventeen HIV reference laboratories, which constitute the national network of HIV-VL and EID testing coverage. For this study, the number of rejected samples, the reason for rejection, and the type of test ordered were recorded monthly.

**Results:** During the study period, of the 326,885 specimens received for HIV viral load, 12,748 (3.9%) were rejected, and 2.7% (1,039) of the 38,354 specimens received for EID were also rejected. The analysis of SNC shows the presence of more than one error or NC in some samples. For HIV viral load, our results indicate that specimen identification errors for viral load were the most common NC (63.14%; n=8049; P=0.031), followed by insufficient specimen volume (43.7%; n=5571; P=0.049) and quality errors, including hemolyzed specimens (27.8%; n=3543; P=0.054), and specimen transport packaging errors (9.1%; n=1160; NS). For HIV EID, specimen rejections were primarily attributed to missing or mismatched identification on the request forms (37.12%, n=386; P=0.042), sample unavailability (13.4%; n=139; P=0.056), and information discrepancies (44.2%; n=459; P=0.033). The observed significant rejection rates for both HIV viral load and EID exceeded the established national rejection rate of <2% of errors. Our results suggest that corrective action is critical, along with the establishment of policies to detect and resolve preanalytical errors in Cameroon.

**Conclusion:** Our findings highlight the high magnitude of preanalytical errors for HIV-VL and EID tests used in the testing and management of people living with HIV/AIDS in Cameroon. Therefore, the laboratory system should be strengthened to provide quality services to patients and support optimization. Suggestions for improvement include the distribution of a validated specimen collection manual, the creation of electronic test request forms, staff training, and regular on-site supervision of the use of available resources is necessary in this country.

## Introduction

The laboratory testing process encompasses preanalytical, analytical, and postanalytical phases (1). Of these three phases, the preanalytical phase spans from the initial test request to the preparation of the sample for analysis. Notably, investigations into laboratory processes have shown that errors predominantly arise during the pre-analytical stage, and errors occurring during this phase account for approximately 70% of all laboratory errors (2,3).

This high error rate can be attributed to the external factors and the manual handling of specimens inherent to this phase, and its effects frequently appear in the pre-analytical phase (4,5). If left unchecked, preanalytical errors can significantly compromise the reliability and accuracy of test results, ultimately affecting patient care and increasing healthcare costs. The frequency of errors in laboratory testing is often linked to the handling of samples, including test requests, specimen collection, storage, transportation, and labeling, which are typically managed externally and performed manually (6,7).

HIV Viral Load (HIV-VL) and Early Infant Diagnosis (EID) play a pivotal role in the laboratory surveillance, monitoring of HIV/AIDS (8,9). The integration of automation, databases, and computer systems has revolutionized laboratory operations, enabling faster and more accurate tests of HIV viral load and EID results. Nevertheless, the preanalytical stage continues to pose significant challenges, as errors or inconsistencies during this phase can have far-reaching and detrimental consequences for patient care (10–12). To provide high-quality laboratory services, precision, accuracy, and timely reporting are essential. Despite significant advancements in laboratory science, errors can still occur due to manual and systemic factors(13). Reducing medical errors not only saves lives but also has a profound social impact on patients and healthcare providers (6,7).

To minimize errors in the pre-analytical phase and achieve high-quality standards, meticulous attention must be paid to this critical stage. Healthcare providers bear the responsibility of ensuring the accuracy and authenticity of patient identification details, which demands utmost diligence to minimize errors (14,14,15). Moreover, healthcare professionals are directly responsible for providing comprehensive information regarding suspected or confirmed diagnoses in a clear, legible, and abbreviation-free manner. This approach not only justifies the relevance of requested tests but also provides laboratory staff with a valuable clinical context, facilitating prompt and informed action (16).

Many HIV reference laboratories have been retained in Cameroon, with more than 15 HIV Viral Load Reference Laboratories and more than 04 HIV Early Infant Diagnosis (17). Most of these HIV reference laboratories are located in the political capital of the country, Yaounde. These reference laboratories provide specialized HIV viral load and EID testing of more than 90% of all specimens from HIV patients on antiretroviral treatment at the national level(18,19). In Cameroon and many countries in the central Africa region, there is a knowledge gap regarding pre-analytical errors during HIV viral load and EID laboratory processes (20). This study aimed to investigate factors contributing to pre-analytical errors in the HIV viral load and early infant reference laboratories in Cameroon to strengthen the laboratory system in order to support surveillance and monitoring of HIV/AIDS patients, as well as support the optimization of the use of available resources.

## Material and Methods

### Study design and setting

A cross-sectional, descriptive, and quantitative study design was conducted from January to December 2024 in all HIV reference laboratories in Cameroon. These laboratories provide a national network of HIV viral load and early infant diagnosis (EID) testing coverage throughout the country. For this study, assessment of pre-analytical errors was conducted in all 17 functional HIV reference laboratories in Cameroon (**Figure 1**).

**Figure 1:**
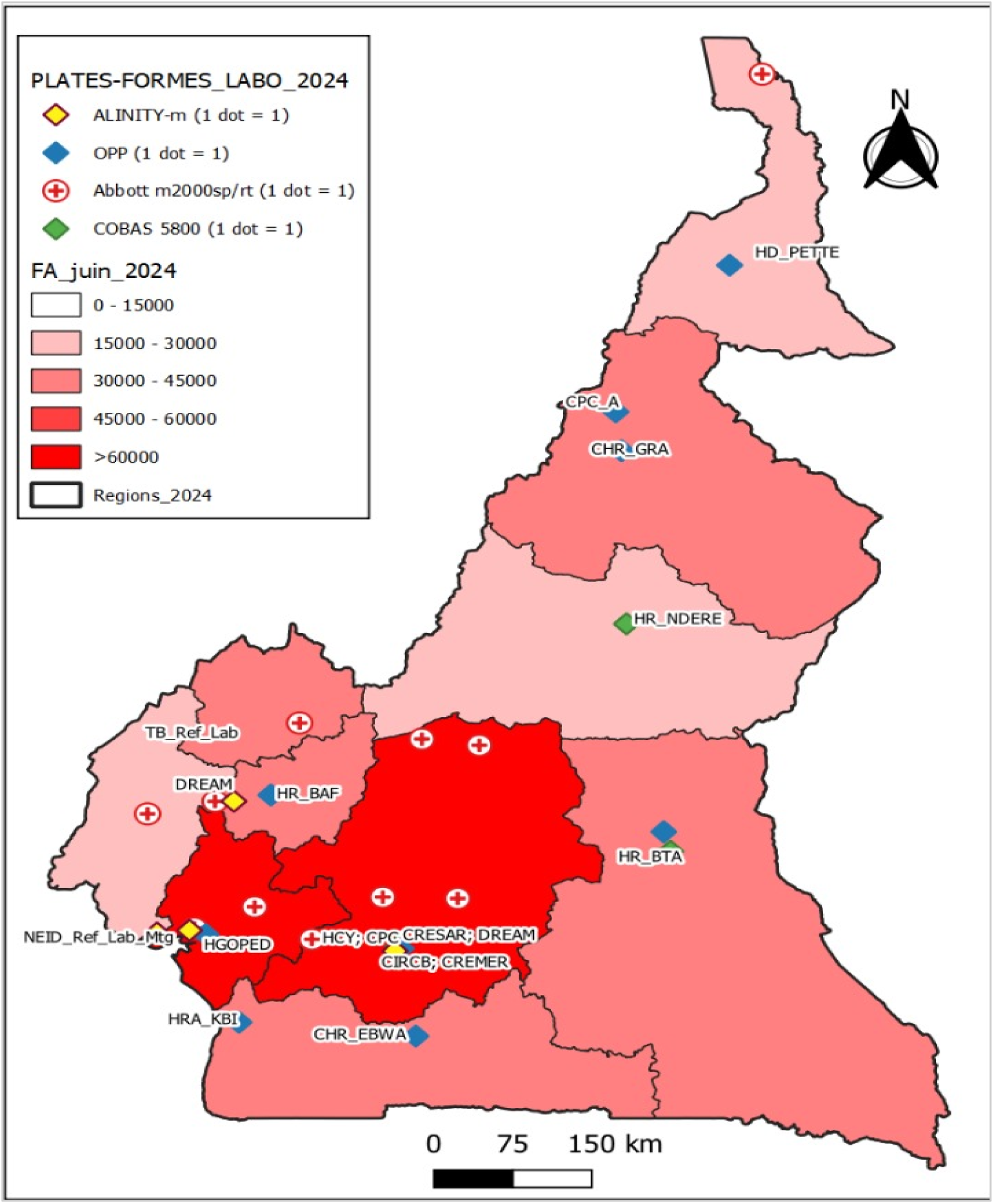
HIV viral load and EID reference laboratories of Cameroon

### Sampling and data collection

For this study, data were collected using self-administered structured standard questionnaires and the harmonized HIV viral load and/or EID specimen collection and transport checklists used for sending HIV specimens to reference laboratories in Cameroon. Briefly, all specimens are collected by trained personnel at the HIV care center of agreement, and all outpatient specimens are received by qualified staff at the reference laboratories along with the appropriate request forms. Upon receipt of specimens, all rejected specimens, the reason for rejection, the type of test ordered (HIV viral load or early infant diagnosis), as well as any preanalytical errors or discrepancies, including hemolyzed specimen, insufficient volume, wrong specimen tube, mismatch between name on specimen and request form, poor temperature preservation, missing specimens, delay in specimen transport, identification number given to two different patients, were recorded in the problem reporting logbook.

### Reasons for sample rejection in reference laboratories

Samples were rejected due to various reasons, including inadequate sample volume, improper labeling with essential patient information, or labeling with a non-permanent marker . Additionally, samples that appeared compromised, such as those that were hemolyzed or exhibited visible signs of degradation, were also rejected. Other reasons for rejection included improper packaging, inappropriate temperature, and inadequate drying of samples for the EID test prior to shipping. Furthermore, samples were rejected if they were not accompanied by a completed patient/test request form.

### Ethical consideration

This assessment was deemed exempt from ethical review because it solely focused on the quality assurance of samples submitted for HIV viral load and Early Infant Diagnosis (EID) testing, without involving human subjects or personally identifiable information.

### Statistical analysis

Statistical analysis of all data entered was performed using the Statistical software IBM SPSS Statistics V.25.0. Data were expressed as mean ± SD, and Categorical data were expressed as percentages, and a p-value of less than 0.05 was considered statistically significant.

## Results

### Distribution of samples attributed in 2024 for HIV viral load and EID testing

In this cross-sectional study, a total of 326,885 specimens were submitted to HIV viral load testing, and 38,354 for EID testing. Of note, all of these samples were outpatient samples for the two analyses (100%). A significant correlation between the number of samples received per region and the number of reference laboratories was observed, with 187,631 (57.4%) of samples submitted in the HIV viral load in the Center region (P=0.044) and 24,830 (64.73%) for EID (P=0.033) (**Table 1**). Remarkably, 34.5% (64,732) of samples analyzed in the center region were collected outside the center region in different types of healthcare centers providing care.

**Table 1:**
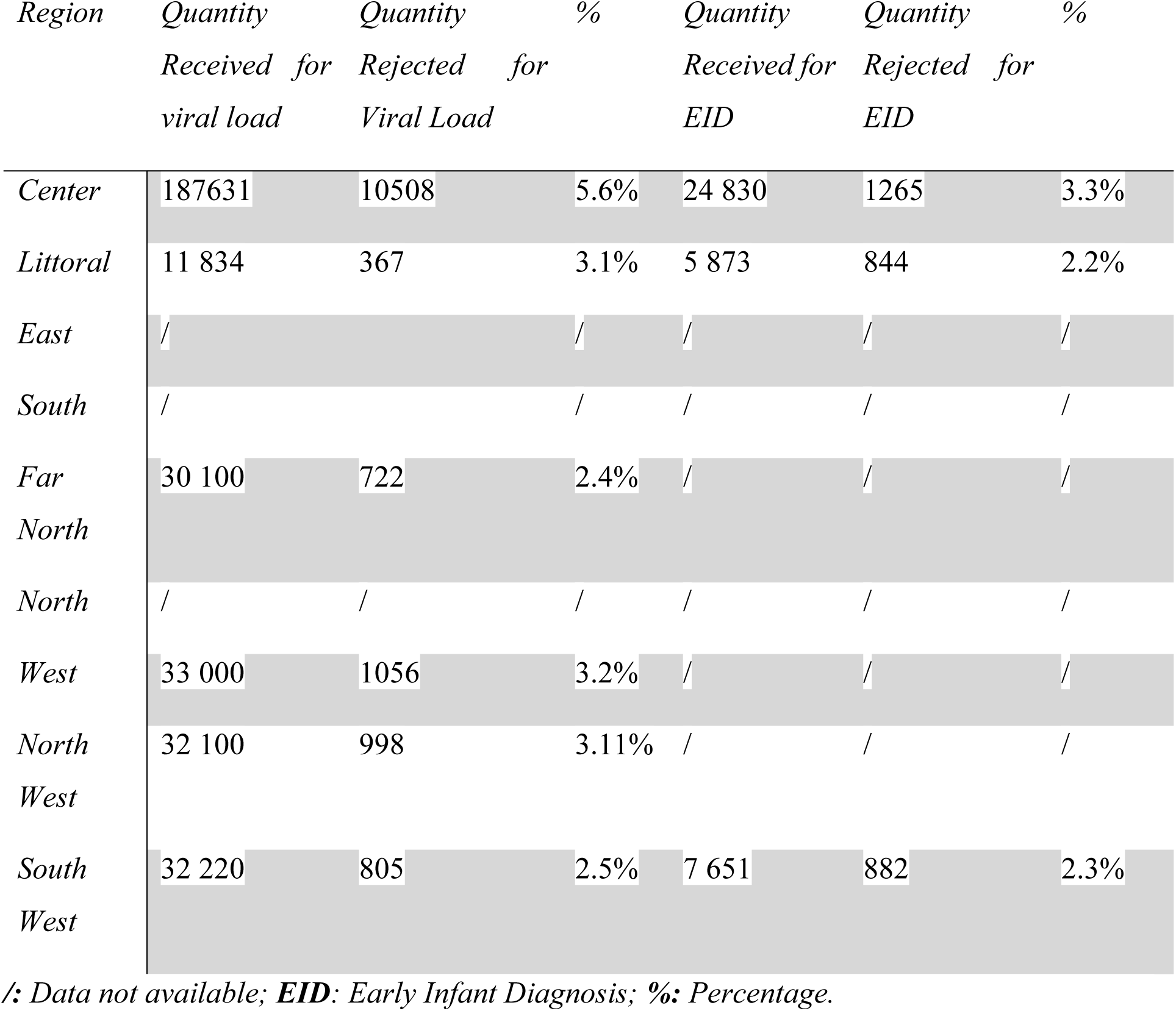
Distribution of samples received and rejected per region attributed for HIV viral load and EID testing in 2024 in Cameroon.

### Distribution of reasons for sample rejection during the pre-analytical stage in HIV viral load and EID testing laboratories

A total of 209 samples of viral load testing and 59 DBS samples for which the reason for rejection was not recorded were excluded from the present evaluation. Of a total number of specimens with the reason of sample rejection, 3.9% (12,740/32,6676) were rejected for HIV viral load testing with the range of rejection value of 2.4% to 5.6%. Regarding samples received for EID analysis, a total of 2.7% (1034/38,295) was rejected, with the rejection interval of 2.2% to 3.3%. For these two analyses, the highest rejection rate was observed in samples collected outside the region where the reference laboratory is located and in health facilities with a high proportion of sample collection. The analysis of the total number of preanalytical errors observed shows the presence of more than one reason for rejection in some samples. In the present study, the most common reason for HIV viral load sample rejection is related to identification (mislabeling or double identification) (63.14%; n=12 748; P=0.031), followed by insufficient sample volume (43.7%; n=5571; P=0.049), and quality errors such as hemolyzed samples (27.79%; n=3541) and packaging errors (9.058%; n=1160). Regarding EID analysis, missing sample and/or request form were predominantly identified, followed by identification information. **(Table 2**).

**Table 2:**
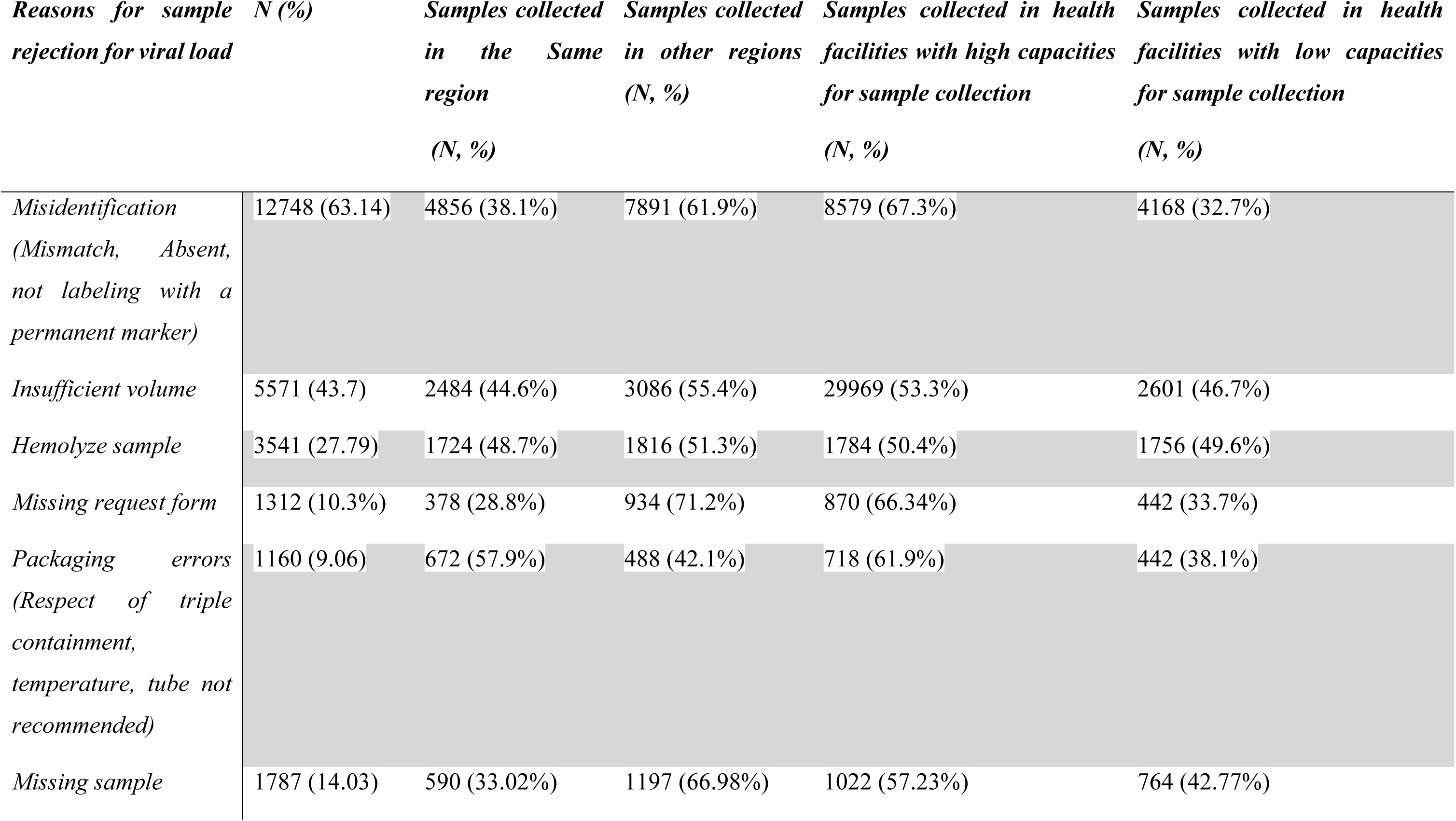
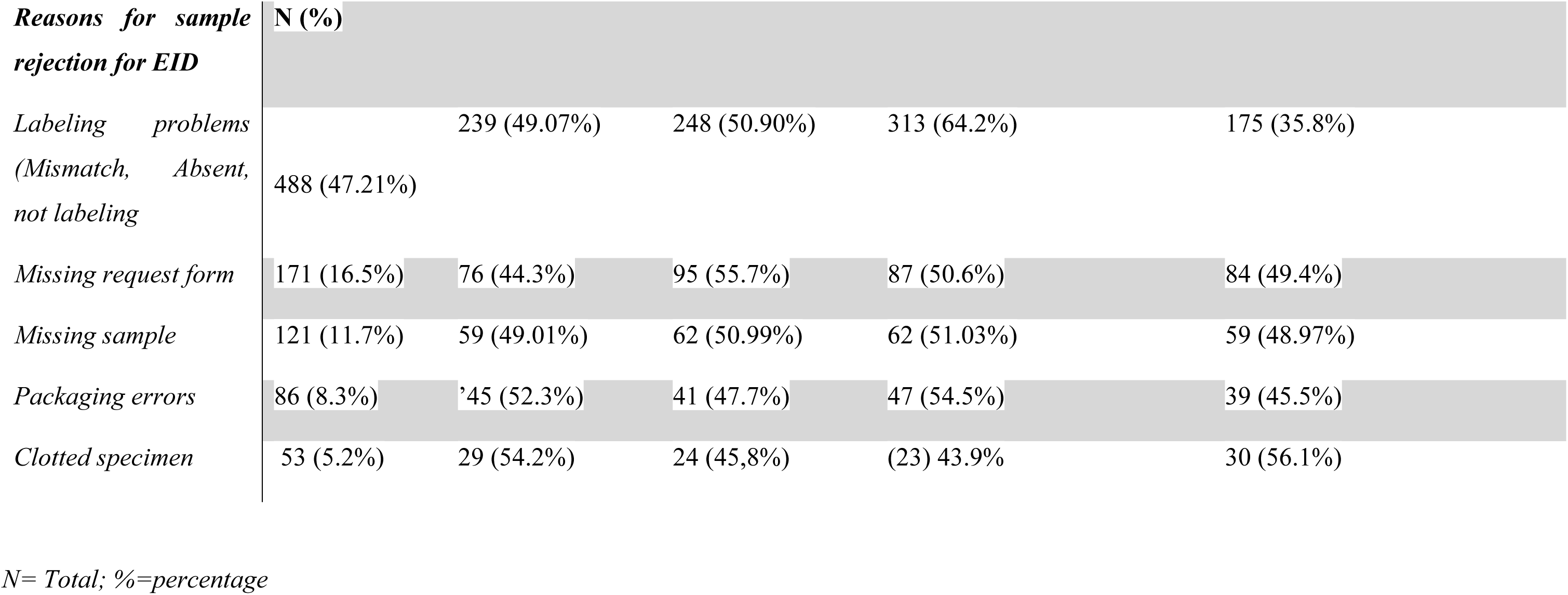
Distribution of reasons for sample rejection during the pre-analytical stage in HIV viral load and EID testing laboratories.

### Factors affecting pre-analytical errors

The analysis of factors affecting pre-analytical errors or non-conformity observe during sample reception shows that identification errors and or mismatch were the most common NC (63.14%; n=8049; P=0.031), followed by insufficient volume of sample (43.7%; n=5571; P=0.049) and quality errors, including hemolyzed samples (27.79%; n=3543), and packaging errors (09.058%; n=1160). Regarding HIV EID, specimen rejection was primarily attributed to missing or incomplete request forms (37.12%, n=386, P=0.042), sample unavailability (13.4%, N=139, P=0.056), and information discrepancies (44.2%, n=459, P=0.033) (**Table 2**).

### Stratification of data by the type of requested analysis and the region or healthcare capacity of the sample collection

Further evaluation was performed to evaluate the contributing factors among the 63.14% (8049/12748) of errors associated with identification or labeling errors in this study for both HIV viral load. Of the total sample for which labeling errors occurred, 44.5% (3581/8049) were not unambiguously identified due to non-permanent marker identification, 21.7% (1746/8049) had double identification, 33.2% (2672/8049) had forgotten identification, and 4.04% (322/8049) were samples present on the checklist but physical samples were not available (**Table 2)**.

Stratification of the data by type of analysis requested, region, and localization of the capacity of the number of patients cared for in the health facility that collected samples was also done. A significant disparity was observed in rejection rates between samples collected from the same region and those from clinics or rural agreement HIV health facilities (P=0.041 and P=0.037), **Table 3**. Our findings revealed that the proportion of preanalytical errors was significantly higher in specimens received from other regions compared with specimens collected in the same region of testing (76.4% vs 23.6%; P=0.022) and (69.1% vs. 30.9%; P=0.033) for HIV viral load and EID, respectively. In addition, the magnitude of errors based on region and localization of the health facility accounts for the higher (55.6% vs 44.4%, n=, P=0.042) for viral load and (50.7% vs 49.3%, n=, P=0.052) for EID. The proportion of rejected samples ranged from 2% to 4% for HIV viral load and 1% to 3% for EID samples, with the highest rejection rate observed for samples collected out of the testing region and health facilities with high capacity for sample collection. Remarkably, samples collected in healthcare facilities where most patients receive care were five times more likely to be rejected than those in facilities that care for a small number of patients (**Table 3**).

**Table 3:** Relationship between sample rejection and the region of sample collection or the capacity of sample collection.

| ***Viral load:***<br>***Samples collected*** | ***Samples received***<br>***(%)*** | ***Sample rejected***<br>***(%)*** | ***Odds ratio (95% CI)*** |
| --- | --- | --- | --- |
| *The Same region* | 257 324 (78.72%) | 6 690 (2.6%) | 2.21 (1.92-5.87) |
| *In other regions* | 69 561 (21.28%) | 3756 (5.4%) | 5.4 (3.79-9.65) |
| *In health facilities*<br>*with high*<br>*capacities for*<br>*sample collection* | 274257 (83.9%) | 13 987 (5.1%) | 4.17 (2.67-9.57) |
| *In health facilities*<br>*with low capacities*<br>*for sample*<br>*collection* | 52628 (16.1%) | 1210 (2.3%) | 2.16 (1.26-7.24) |
| ***EID Samples***<br>***collected*** |  |  |  |
| *The Same region* | 789 (76.2%) | 19 (2.4%) | 2.32 (1.97-5.80) |
| *In other regions* | 246 (23.8%) | 8 (3.2%) | 4.24 (2.58-11.12) |
| *In health facilities*<br>*with high*<br>*capacities for*<br>*sample collection* | 862 (83.4%) | 30 (3.5%) | 5.12 (3.72-10.31) |
| *In health facilities*<br>*with low capacities*<br>*for sample*<br>*collection* | 175 (16.9%) | 4 (2.2%) | 2.23 (1.91-9.63) |

## Discussion

Despite the advent of automation in HIV viral load and EID laboratories and efforts to improve HIV molecular testing, analytical quality errors continue to occur throughout the laboratory testing cycle (1,2,21). It is well known that enhancing the quality of the results of the laboratory necessitates a proactive approach to identifying, recording, and tracking errors that can undermine the reliability of test results (4–6).

This cross-sectional study, based on HIV reference laboratories, investigated for the first time the common reasons for sample rejection and identified factors influencing pre-analytical errors in the national network of HIV VL and EID testing coverage in Cameroon. The country’s HIV viral load and EID national guidelines have fixed the maximum range of target rejection rate at less than 2% per month (22). Our findings indicate that the HIV viral load or EID rejection rate surpasses this threshold (3.9% and 2.7%, respectively), highlighting the need for targeted interventions to optimize sample quality and reduce rejection rates due to pre-analytical errors resulting from poor management of samples in health facility responsible for the biological monitoring of HIV infected patients in Cameroon. The rejection rate found in the present study is relatively high for HIV viral load and similar to that found in studies conducted in Ethiopia, 3.6% (4), and the literature were several studies have documented high prevalence errors during pre-analytical stage in the laboratory (4,5,23). Our results, however, contrast those of the EID rate of rejection in Zimbabwe, 2 to 7% depending on the region (5). The high incidence of specimen rejection in the present study underscores the need for preventive strategies to minimize the risks associated with incorrect or inadequate specimen collection, including delayed results, diagnostic errors, adverse medication events, and compromised patient safety. In addition, the significant proportion of rejected specimens in the present study highlights the need for preventive measures to address the issues arising from suboptimal specimen collection, such as delayed results, incorrect diagnoses, and adverse patient outcomes.

The current study found that the most common pre-analytical non-conformities or rejections in HIV viral load or EID testing were sample identification or Mismatched errors, insufficient volume, and hemolyzed samples. Our findings are consistent with the results of the studies conducted in Ethiopia (23), in Zimbabwe (5), and the literature (4,23–25). Our results are different from the findings of another study conducted in Ethiopia, where using an inappropriate sample collection container was the predominant reason for sample rejection (4).

In addition, this study showed that samples collected at different regions, different from the region of the HIV viral load and or EID testing, were four times more likely to be rejected. Our results are similar to the studies conducted in Nigeria (26) and Zimbabwe (5). Remarkably, samples collected in healthcare facilities where most patients receive care were five times more likely to be rejected than healthcare facilities that received a small number of patients in care. Findings are similar to a Nigerian study, where the bulk of rejected samples came from facilities that serve most patients (26).

## Limitations of the Study

Interviewed results were dependent on the laboratory personnel’s response. So, biased information might be given as data were collected during their daily working time. However, the observational data helps to minimize this bias. Regarding both HIV viral load and EID, there were no available data to track the interval between sample collection and informing the mother or HIV patient that the sample had been rejected. Such data could show whether sample rejection adversely influences early ART initiation for infants.

## Conclusion

Despite significant advancements in the analytical phase of clinical laboratory testing, errors persist, particularly in the pre-analytical phase. Our study reveals a substantial prevalence of pre-analytical errors in HIV-VL and EID tests, which are crucial for diagnosing and managing HIV/AIDS patients in Cameroon. To address this issue, the laboratory system requires strengthening to ensure high-quality services and optimal resource utilization. Recommendations for improvement include implementing a validated sampling manual, introducing electronic test request forms, providing staff training, and conducting regular on-site supervision.

## Data Availability

All relevant data are within the manuscript and its Supporting Information files.

## Acknowledgments

We would like to thank all the staff of the HIV reference laboratories for their involvement in quality control records in this study.

## Funding sources

The author(s) received no specific funding for this work.

## Competing interests

All the authors declare that they have no competing interests.

## Supporting Information

S1 Table 1: **Distribution of samples received and rejected per region attributed for HIV viral load and EID testing in 2024 in Cameroon**.

S2 Table 2: **Distribution of reasons for sample rejection during the pre-analytical stage in HIV viral load and EID testing laboratories**.

S3 Table 3: **Relationship between sample rejection and the region of sample collection or the capacity of sample collection**.

## Notes

### Competing Interest Statement

The authors have declared no competing interest.

## References

1. Martins JM, Rateke ECM, Martinello F. Assessment of the pre-analytical phase of a clinical analysis laboratory. J Bras Patol E Med Lab. 2018 Aug;54:232–40.

2. Iqbal MS, Tabassum A, Arbaeen AF, Qasem AH, Elshemi AG, Almasmoum H. Preanalytical Errors in a Hematology Laboratory: An Experience from a Tertiary Care Center. Diagnostics. 2023 Jan;13(4):591.

3. Zhang ZH, Wu CC, Chen XW, Li X, Li J, Lu MJ. Genetic variation of hepatitis B virus and its significance for pathogenesis. World J Gastroenterol. 2016 Jan 7;22(1):126–44.

4. Berhan A, Almaw A, Damtie S, Solomon Y, Legese B, Getie B, et al. Determining the rate and reasons for specimen rejection among specimens referred for HIV viral load determination through referral linkage to Debre Tabor Comprehensive Specialized Hospital, north-central, Ethiopia,2023: Retrospective study. Heliyon. 2024 May 22;10(11):e31736.

5. Chiku C, Zolfo M, Senkoro M, Mabhala M, Tweya H, Musasa P, et al. Common causes of EID sample rejection in Zimbabwe and how to mitigate them. PLoS ONE. 2019 Aug 8;14(8):e0210136.

6. Kani V, K K, Sonti S. Assessment of Pre-analytical Errors and Fostering Strategies to Enhance Accurate Results and Efficient Turnaround Times in the Cytology Laboratory of a Tertiary Care Hospital. Cureus. 2024 Mar;16(3):e56592.

7. Nordin N, Ab Rahim SN, Wan Omar WFA, Zulkarnain S, Sinha S, Kumar S, et al. Preanalytical Errors in Clinical Laboratory Testing at a Glance: Source and Control Measures. Cureus. 16(3):e57243.

8. Kamble S, Gawde N, Goel N, Thorwat M, Nikhare K, Bembalkar S, et al. Access, timeliness and retention for HIV testing under early infant diagnosis (EID) program, India. Sci Rep. 2023 Apr 6;13(1):5638.

9. Kyere GA, Vechey GA, Charles-Unadike VO, Tarkang EE. Trends in viral load suppression among HIV patients on antiretroviral therapy (ART) at Asante Mampong Municipal Hospital, Ghana: 2019–2023. BMC Infect Dis. 2024 Oct 16;24(1):1170.

10. Kirk M, Assoa PH, Iiams-Hauser C, Kouabenan YR, Antilla J, Steele-Lane C, et al. Adaptation of an electronic dashboard to monitor HIV viral load testing in Côte d’Ivoire. Afr J Lab Med. 2021 May 17;10(1):6.

11. Parekh BS, Ou CY, Fonjungo PN, Kalou MB, Rottinghaus E, Puren A, et al. Diagnosis of Human Immunodeficiency Virus Infection. Clin Microbiol Rev. 2018 Nov 28;32(1):e00064–18.

12. Drain PK, Dorward J, Bender A, Lillis L, Marinucci F, Sacks J, et al. Point-of-Care HIV Viral Load Testing: an Essential Tool for a Sustainable Global HIV/AIDS Response. Clin Microbiol Rev. 2019 May 15;32(3):10.1128/cmr.00097-18.

13. Balogh EP, Miller BT, Ball JR, Care C on DE in H, Services B on HC, Medicine I of, et al. The Diagnostic Process. In: Improving Diagnosis in Health Care [Internet]. National Academies Press (US); 2015 [cited 2025 Apr 14]. Available from: https://www.ncbi.nlm.nih.gov/books/NBK338593/

14. Salar A, Kiani F, Rezaee N. Preventing the medication errors in hospitals: A qualitative study. Int J Afr Nurs Sci. 2020 Jan 1;13:100235.

15. Riplinger L, Piera-Jiménez J, Dooling JP. Patient Identification Techniques – Approaches, Implications, and Findings. Yearb Med Inform. 2020 Aug;29(1):81–6.

16. Demsash AW, Kassie SY, Dubale AT, Chereka AA, Ngusie HS, Hunde MK, et al. Health professionals’ routine practice documentation and its associated factors in a resource-limited setting: a cross-sectional study. BMJ Health Care Inform. 2023 Feb 16;30(1):e100699.

17. CDC In Cameroon: 20 years of Public Health Impact.

18. Tayong GFE, Sander M, Vuchas C, Samje M, Kum V, Enokbonong P, et al. HIV viral load suppression rates among adults and children living with HIV in the North West Region of Cameroon: A call for action! PLOS ONE. 2025 Jan 31;20(1):e0316399.

19. Bekolo CE, Ndeso SA, Moifo LL, Mangala N, Ateudjieu J, Kouanfack C, et al. Changes in access to viral load testing, incidence rates of viral load suppression and rebound following the introduction of the ‘universal test and treat’ guidelines in Cameroon: A retrospective follow-up analysis. PLOS Glob Public Health. 2024 Apr 16;4(4):e0003042.

20. Kamgaing RS, Bouba Y, Sosso SM, Gabisa JE, Nanfack A, Fokam J, et al. Challenges faced by the HIV testing system in low- and middle-income countries. Afr J Lab Med. 2023 Jan 31;12(1):1974.

21. Zhang L, Hu ZD. Clinical applications of machine learning in pre-analytical, analytical and post-analytical phases of laboratory medicine: a narrative review. AME Med J [Internet]. 2022 Dec 30 [cited 2025 Apr 5];7(0). Available from: https://amj.amegroups.org/article/view/7424

22. Cameroon-Strategic-Direction-Summary-2023.pdf [Internet]. [cited 2025 Apr 7]. Available from: https://www.prepwatch.org/wp-content/uploads/2024/06/Cameroon-Strategic-Direction-Summary-2023.pdf

23. As M. Pre Analytical Errors in the HIV Anti Retro Viral Therapy (ART) Laboratory of Teaching Referral Hospitals in Addis Ababa, Ethiopia. [cited 2025 Apr 4]; Available from: https://www.clinmedjournals.org/articles/ijva/international-journal-of-virology-and-aids-ijva-6-057.php?jid=ijva

24. Dikmen ZG, Pinar A, Akbiyik F. Specimen rejection in laboratory medicine: Necessary for patient safety? Biochem Medica. 2015 Oct 15;25(3):377–85.

25. Getawa S, Aynalem M, Melku M, Adane T. Blood specimen rejection rate in clinical laboratory: A systematic review and meta-analysis. Pract Lab Med. 2022 Dec 19;33:e00303.

26. Auchi Inalegwu SP, Le Abimiku PD. Active tracking of rejected dried blood samples in a large program in Nigeria. World J Virol. 2016 May 12;5(2):73–81.

